# SARS-CoV-2 infection: the environmental endurance of the virus can be influenced by the increase of temperature

**DOI:** 10.1101/2020.05.30.20099143

**Authors:** Fabio Magurano, Melissa Baggieri, Antonella Marchi, Giovanni Rezza, Loredana Nicoletti, COVID-19 Study Group

## Abstract

The COVID-19 disease, a respiratory disease transmitted by a new betacoronavirus SARS-CoV-2. As for other viral respiratory agents, SARS-CoV-2 spreads by person to person through respiratory droplets and direct contact and potentially by indirect contact through fomites. The goal of the current study is to evaluate whether the increase of temperature can influence the environmental endurance of SARS-CoV-2.

We tested SARS-CoV-2 environmental stability in parallel at room temperature (RT, 20°C-25°C) and at average maximum temperature of June (JT) estimated at 28°C in Italy. The virus inoculated on plastic surface was harvested at predefined time-points and tested to evaluate viral titres on Vero cells by TCID50.

Our results confirm that fomite transmission of the emerging SARS-CoV2 is possible, since the virus remains viable on surfaces up to 84 hours at both RT and JT. Moreover, a remarkable difference between the two temperatures exists, suggesting that virus vitality can be influenced by the environmental temperature.

Our results support the hypothesis that in the hot season the increase of temperature could influence the environmental endurance of SARS-CoV2 and reduce Covid-19 transmission probability.

## OBJECTIVES

Corona VIrus Disease 19 (COVID-19) outbreak was declared a pandemic on March 11, 2020. COVID-19 is caused by the enveloped betacoronavirus SARS-CoV-2, transmitted person to person through respiratory droplets and direct contact, and potentially by indirect contact through fomites (1).

Clinically, patients with COVID-19 present with respiratory symptoms, which is very similar to the presentation of other respiratory virus infections. Radiologically, COVID-19 is characterized by multifocal ground-glass opacities, even for patients with mild disease. The case-fatality rate of this disease has been calculated as 7% (2).

As of 17 May 2020, 4,534,731 cases of COVID-19 have been reported globally, including 307,537 deaths. In Europe, 1,870,545 cases have been reported, with United Kingdom, Spain, Italy, Germany and France as the five most reporting countries. In Italy, 224,760 cases were reported and 31,763 deaths, with a 14.1% mortality rate higher rates among elderly patients and patients with comorbidities.

Lockdown restrictions have been introduced in many countries in response to the spread of SARS-CoV-2 to control this emerging global health threat. In Italy, the lockdown started on March 9, 2020, when Italian citizens were strictly encouraged to take preventive measures and maintain social distancing. The lockdown measures have permitted hospitals and medical centers successfully handling patients flow to local hospitals, and addressing issues about hospital beds, overcrowding in emergency departments, and patients transfer to other specialized facilities. All the national administrators of websites have encouraged companies to offer free online services, and educational institutions and universities adopted e-learning methods. Moreover, any data or updates on COVID-19 have been made available for free to general public. As of 4 May 2020, Italy entered phase two of the COVID-19 emergency, with the start of the gradual relaxation of the lockdown measures, but the contagion risk remains high.

Several studies have shown that viral spread could be influenced by climatic conditions since enveloped viruses tend to reduce their circulation in summertime owing to high temperature and solar radiation (3, 4). No wonder, current spread of COVID-19 along the equator and tropics has been significantly less (5). The world is waiting the summer with the hope that the increase of temperature will influence the environmental endurance of SARS-CoV-2.

In Italy, phase two of the emergency is close to the summer, and the increase of temperature may reduce COVID-19 prevalence. In order to try to predict the effect on the epidemic dynamic of COVID-19 during the next months, we decided to test SARS-CoV-2 environmental stability in parallel at room temperature (RT, 20°C-25°C) and at average maximum temperature of June (JT) estimated at 28°C in Italy.

## METHODS

The strain *BetaCov/Italy/CDG1/2020IEPI ISL 41297312020-02-20* (6) was used to test SARS-Cov-2 stability on plastic surface (polypropylene).

The viral preparation was spotted in droplets of 10 µl on 24-well plates and let 30 minutes to dry. Then, plates were incubated at both RT and JT for 7 days at relative humidity of 35–45% (airconditioned room). The virus was harvested at t= 0, 4, 8, 12, 24, 36, 48, 60, 72, 84, 96 hours by adding 1 ml/well of MEM + 2% Fetal Calf Serum (FCS) and quantified by end-point titration on Vero cells by TCID_50_,. In brief, 96-well plates were inoculated with two-fold dilutions of each viral sample (100 µl), and 22,000 cells/well in MEM + 2% FCS were added (100 µl). Plates were left in incubator at 37°C with 5% CO_2_ for 6 days and checked daily to observe the cytopathic effect (CPE). The 50% endpoint titers have been determinate using the Spearman-Karber method. Both the experiments were conducted in three independent replicates.

Viral titre of the daily collections (0, 24, 48, 72 hours) at RT were determined also by plaque assay in Vero E6 cells. Briefly, 12-well plates were plated with Vero E6 cells (150,000/well in MEM +10% FCS) and inoculated with logarithmic dilutions of each sample. Plates were incubated 1 hour at 37°C, and 4 ml/well of a medium containing 2% Gum Tragacanth + MEM 2.5% FCS were added. After 5 days at 37°C with 5% CO2, titres were calculated by crystal violet dyeing in plaque-forming units per milliliter (PFU/ml).

Standard deviation (SD) of three wells at different time from 0 to 84 hours was performed in order to visualize the dispersion of a dataset relative to its mean.

## RESULTS

The strain *BetaCov/Italy/CDG1/2020|EPIISL 412973|2020-02-20* used to test SARS-Cov-2 stability in vitro had an initial viral titre of 10^6.8^ TCID_50_/ml, a comparable viral load of symptomatic, asymptomatic or minimally symptomatic patients (7). Contaminated surfaces are known to be significant vectors in the transmission of infections and survival of viruses on a variety of fomites has been demonstrated for influenza viruses, paramyxoviruses, poxviruses, and retroviruses (8, 9). Our study confirms that fomite transmission of the emerging SARS-CoV-2 is possible (10), since the virus remains viable on plastic surfaces up to 84 hours at both RT and JT. In both the experimental conditions, the virus is not detectable anymore at 96 hours.

Results by end-point titration show that the virus vitality on plastic surface rapidly declined by more than 1 log10 in TCID_50_ during the first 24-36 hours, at RT. This trend is confirmed by titration by plaque assay (Appendix). At JT the same decay was observed more rapidly (between 8 and 12 hours), indicating that viral infectivity can be influenced by higher temperature. A remarkable difference between the two temperatures exists, suggesting that virus vitality can be influenced by the environmental temperature (Figure). In this study, 95% of confidence interval (CI) was considered statistically significant (Appendix).

**Figure.**
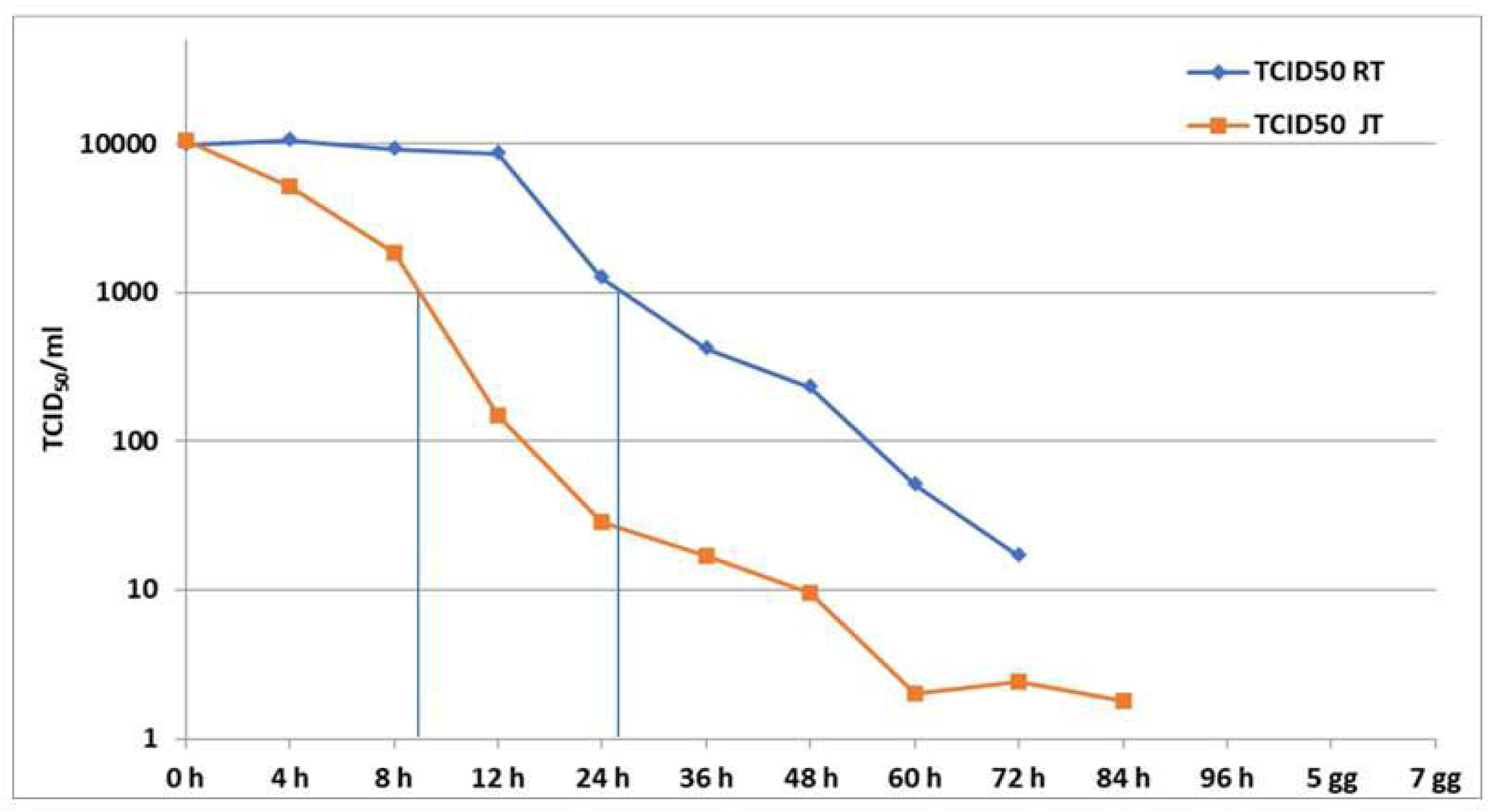
Relation between viral titre (TCID50) and time of collection in logaritmic scale.

**Figure 1.**
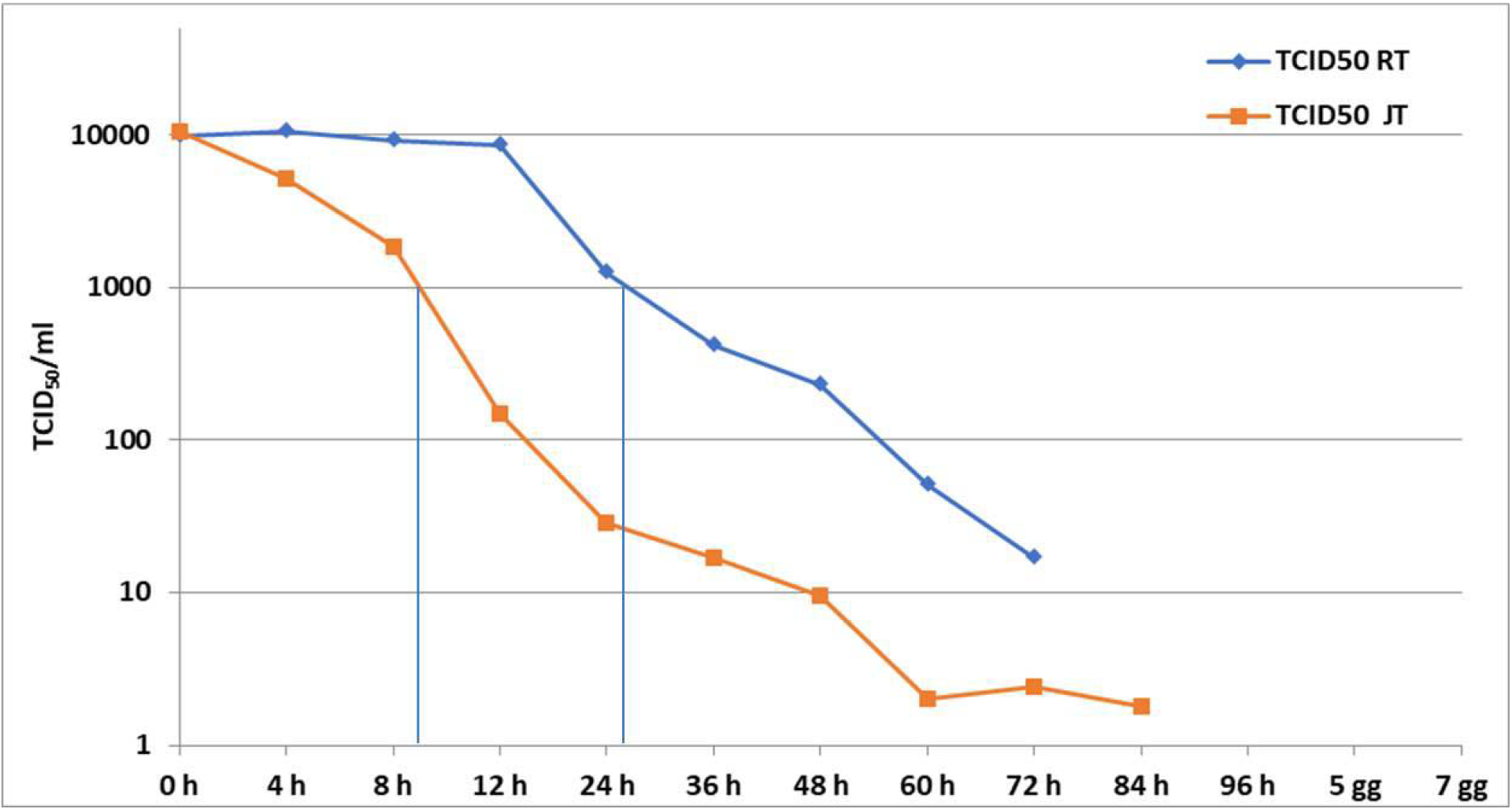
Relation between viral titre (TCID_50_) and tine or collection in logaritmic scale. Average viral titers in samples collected at either RT or JT are plotted as function of time-points post-infection. At RT. an important reduction in the viral titre. from 4 log 10 to 3 log10 TCID_50_. per milliliter of medium, was observed during the first 24-36 hours. At JT the same decay was observed more rapidly (between 8 and 12 hours) indicating that viral inactivity can be influenced by higher temperature. This dacay trend showing a remarkable difference between the two temperatures continues until 84 hours. In both the experimental conditions, the virus is not detectable anymore at 96 hours.

## CONCLUSIONS

The results of this study support the hypothesis that in the hot season the increase of temperature may influence the environmental endurance of the SARS-CoV-2 and reduce COVID-19 transmission probability. Accordingly, we can assume that the incoming summer might help decline of the number of infected people limiting the risk of new outbreaks of Covid-19.

However, these results should be interpreted with caution and do not influence the need of maintaining social distancing measures.

## Data Availability

NOT AVAILABLE

## Acknowledgements

The authors wish to thank Eugenio Sorrentino, Alessia Caratelli, Ambrogio Carlei, Marina Sbattella for their technical support.

## Authors’ contributions

M. F. conceived and designed the study, M. F., B. M. and M. A. performed the experiments, M. F., B. M. and N. L. analysed the data, M. F., B. M. wrote the manuscript. N. L. and R.G. critically revised the manuscript.

